# SARS-CoV-2 lineage dynamics in England from September to November 2021: high diversity of Delta sub-lineages and increased transmissibility of AY.4.2

**DOI:** 10.1101/2021.12.17.21267925

**Authors:** Oliver Eales, Andrew J. Page, Leonardo de Oliveira Martins, Haowei Wang, Barbara Bodinier, David Haw, Jakob Jonnerby, Christina Atchison, The COVID-19 Genomics UK (COG-UK) Consortium, Deborah Ashby, Wendy Barclay, Graham Taylor, Graham Cooke, Helen Ward, Ara Darzi, Steven Riley, Marc Chadeau-Hyam, Christl A. Donnelly, Paul Elliott

**Author notes:** Corresponding authors: Paul Elliott and Christl A Donnelly, School of Public Health, Imperial College London, Norfolk Place, London, W2 1PG. https://www.cogconsortium.uk (The COVID-19 Genomics UK (COG-UK) Consortium). Full list of consortium names and affiliations is provided as a supporting document (The COVID-19 Genomics UK (COG-UK) Consortium).

## Abstract

Since the emergence of SARS-CoV-2, evolutionary pressure has driven large increases in the transmissibility of the virus. However, with increasing levels of immunity through vaccination and natural infection the evolutionary pressure will switch towards immune escape. Here we present phylogenetic relationships and lineage dynamics within England (a country with high levels of immunity), as inferred from a random community sample of individuals who provided a self-administered throat and nose swab for rt-PCR testing as part of the REal-time Assessment of Community Transmission-1 (REACT-1) study. From 9 to 27 September 2021 (round 14) and 19 October to 5 November 2021 (round 15), all lineages sequenced within REACT-1 were Delta or a Delta sub-lineage with 44 unique lineages identified. The proportion of the original Delta variant (B.1.617.2) was found to be increasing between September and November 2021, which may reflect an increasing number of sub-lineages which have yet to be identified. The proportion of B.1.617.2 was greatest in London, which was further identified as a region with an increased level of genetic diversity. The Delta sub-lineage AY.4.2 was found to be robustly increasing in proportion, with a reproduction number 15% (8%, 23%) greater than its parent and most prevalent lineage, AY.4. Both AY.4.2 and AY.4 were found to be geographically clustered in September but this was no longer the case by late October/early November, with only the lineage AY.6 exhibiting clustering towards the South of England. Though no difference in the viral load based on cycle threshold (Ct) values was identified, a lower proportion of those infected with AY.4.2 had symptoms for which testing is usually recommend (loss or change of sense of taste, loss or change of sense of smell, new persistent cough, fever), compared to AY.4 (p = 0.026). The evolutionary rate of SARS-CoV-2, as measured by the mutation rate, was found to be slowing down during the study period, with AY.4.2 further found to have a reduced mutation rate relative to AY.4. As SARS-CoV-2 moves towards endemicity and new variants emerge, genomic data obtained from random community samples can augment routine surveillance data without the potential biases introduced due to higher sampling rates of symptomatic individuals.

## Introduction

Since its first documented case in India in November 2020 [1] the Delta variant of SARS-CoV-2 has spread rapidly across the world and by 16 November 2021 was responsible for 99.7% of all SARS-CoV-2 infections [2]. Its rapid rise to dominance has been attributed to greater levels of transmissibility [4,5] than previously circulating variants with the reproduction number estimated to be over two-fold higher [6], as well as possible reduced vaccine effectiveness against infection [3]. Since its global dissemination, continued adaptive evolution has led to a diverse set of Delta sub-lineages, with distinct combinations of mutations (especially on the spike protein) [7,8].

Since July 2021 the lineage AY.4.2 (Pango nomenclature [9]), a descendant of the original Delta variant (henceforth B.1.617.2) has increased in proportion in routine surveillance data for England from 8.5% the week beginning 4 October [10] to 14.7% the week beginning 31 October [11]. AY.4.2 was declared a variant under investigation (VUI) by the UK Health Security Agency on 20 October 2021 [12]. Globally AY.4.2 had been detected in 43 countries by 22 November 2021 [13] but had only been estimated at a cumulative proportion greater than 1% in Poland [14]. AY.4.2 has two defining mutations, Y145H and A222V, but is otherwise similar to AY.4, a lineage that is far more widespread. AY.4 is the most prevalent lineage in England (on 29 October 2021) [11] and has been detected in 87 countries (by 22 November 2021) [15], in some of which it had already been reported as the most prevalent lineage (by 23 November 2021) [16,17].

England has recorded high levels of SARS-CoV-2 infection over the course of the pandemic [5,18] and vaccinated a large proportion of its population (80.3% of over 12 year olds double vaccinated by 27 November 2021), with further booster jabs being rolled out in adults (30.5% of over 12 year olds having received a booster dose by 27 November 2021) [18]. This has led to high levels of antibodies against coronavirus with 92.8% of adults in England estimated to test positive for antibodies in the week beginning 1 November 2021 [19]. With high vaccination coverage in the population it is likely that selective pressure on SARS-CoV-2 has shifted towards immune escape. Genomic surveillance in highly immunised regions is crucial to detect emerging variants that can more successfully navigate the immune landscape that has been created by both natural infection and vaccination.

The REal-time Assessment of Community Transmission - 1 (REACT-1) study is a series of cross-sectional surveys of the population of England that seeks to estimate the prevalence of SARS-CoV-2 on a monthly basis [5,20], with genomic sequencing performed on all positive samples with a low enough cycle threshold (Ct) value (a proxy for viral load) and high enough volume. Due to its sampling procedure it does not suffer from the biases of routine surveillance that can be heavily biased towards symptomatic individuals [21]; symptom status can be highly dependent on levels of immunity [22]. Here we present the genomic analysis of the (N=2163) positive samples for round 14 and round 15 which were collected from 9 September to 27 September 2021 and 19 October to 5 November 2021 respectively.

## Results

### Lineage diversity

In round 14 the lineage was determined for 481 of 764 positive samples. All lineages were Delta or a Delta sub-lineage with the four most prevalent lineages being AY.4 at 65.1% (60.7%, 69.2%, n=313), AY.43 at 6.0% (4.2%, 8.5%, n=29), B.1.617.2 (original Delta variant) at 5.2% (3.6%, 7.6%, n=25) and AY.4.2 at 4.6% (3.0%, 6.8%, n=22) (Figure 1-A, Supplementary Table 1). In round 15 the lineage was determined for 840 of 1399 positive samples. Again all samples were Delta or a Delta sub-lineage with the most prevalent lineages again being AY.4 at 57.6% (54.2%, 60.9%, n=484), B.1.617.2 at 12.8% (10.8%, 15.3%, n=108), AY.4.2 at 11.8% (9.8%, 14.1%, n=99) and AY.43 at 4.8% (3.5%, 6.4%, n=40).

**Figure 1:**
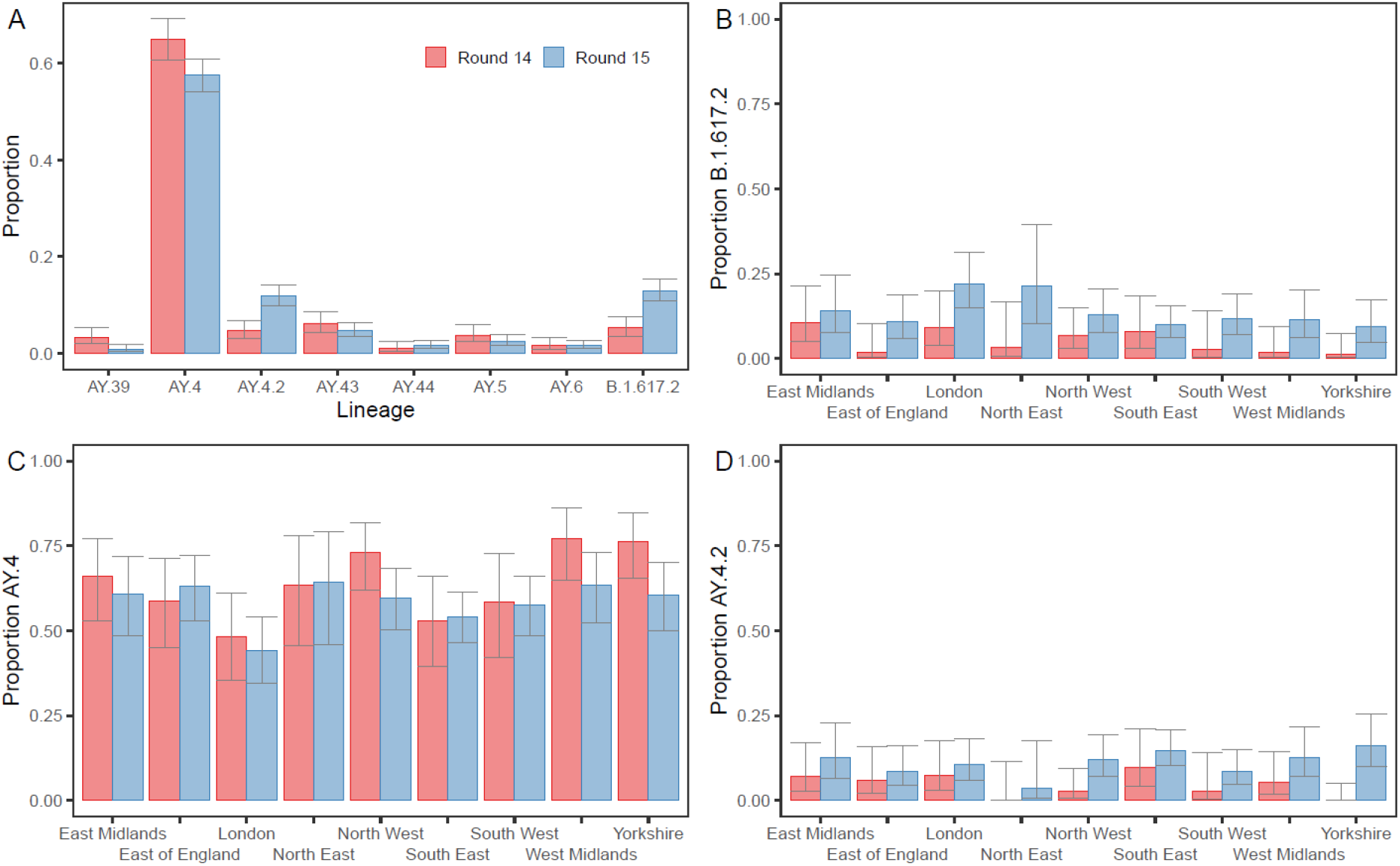
Proportion of positives by Delta sub-lineage. (A) The proportion of positives samples by round designated to the 8 lineages most prevalent over both rounds 14 and 15 (AY.39, AY.4, AY.4.2, AY.43, AY.44, AY.5, AY.6, B.1.617.2). (B-D) Proportion of positive samples by round and region with lineage designated as (B) B.1.617.2, (C) AY.4 and (D) AY.4.2.

The next four most prevalent lineages over both rounds combined were AY.5, AY.6, AY.39, and AY.44. However, even a single detection of a lineage corresponded to an average of 971 (157, 5852) swab-positive individuals at any time during round 14 and 1051 (175, 6264) swab-positive individuals at any time during round 15. During rounds 14 and 15 there were 33 and 31 unique lineages detected, respectively with 44 unique lineages detected overall. There was no apparent difference in genetic diversity between the two rounds as estimated by the Shannon diversity (p = 0.831) (Supplementary Table 2).

### Distribution by region and age

During round 15 the proportion of B.1.617.2 was found to be highest in London at 22.1% (14.9%, 31.4%), being greater than the proportion in South East, East of England and Yorkshire and The Humber (Figure 1-B, Supplementary Table 3). Conversely, in round 14 and 15 the proportion of AY.4 was lowest in London at 48.1% (35.4%, 61.1%) and 44.2% (34.6%, 54.2%) respectively and was found to be higher in North West, West Midlands and Yorkshire and The Humber during both rounds (Figure 1-C, Supplementary Table 3). This reduced proportion of the nationally most prevalent lineage (AY.4) in London coincided with a higher level of genetic diversity in London. The Shannon diversity was highest in London during both rounds at 1.814 in round 14 and 1.809 in round 15 (p <0.001 and p = 0.002 respectively, reference = West Midlands, Supplementary Table 2). Higher levels of genetic diversity were also found during both rounds in the South East and South West, relative to the West Midlands (which showed the lowest levels of genetic diversity in round 14 and the second lowest in round 15). There were no regional differences in the proportion of AY.4.2 during round 15 (Figure 1-D, Supplementary Table 3). Regional differences during round 14 and regional differences for other lineages could not be investigated due to small sample sizes but numbers are provided in Supplementary Table 4.

Sub-regional analysis was performed in order to investigate the presence of clustering in each round for each lineage (see Methods). Despite being highly geographically dispersed (Figure 2) clustering was detected in round 14 for AY.4 (p = 0.037) and AY.4.2 (p = 0.029) (Supplementary Table 5). However, during round 15 clustering was no longer evident for both AY.4 (p = 0.706) and AY.4.2 (p = 0.067). The only lineage for which clustering was detected in round 15 was AY.6 (p = 0.003) which was found mainly in London and towards the South coast of England.

**Figure 2:**
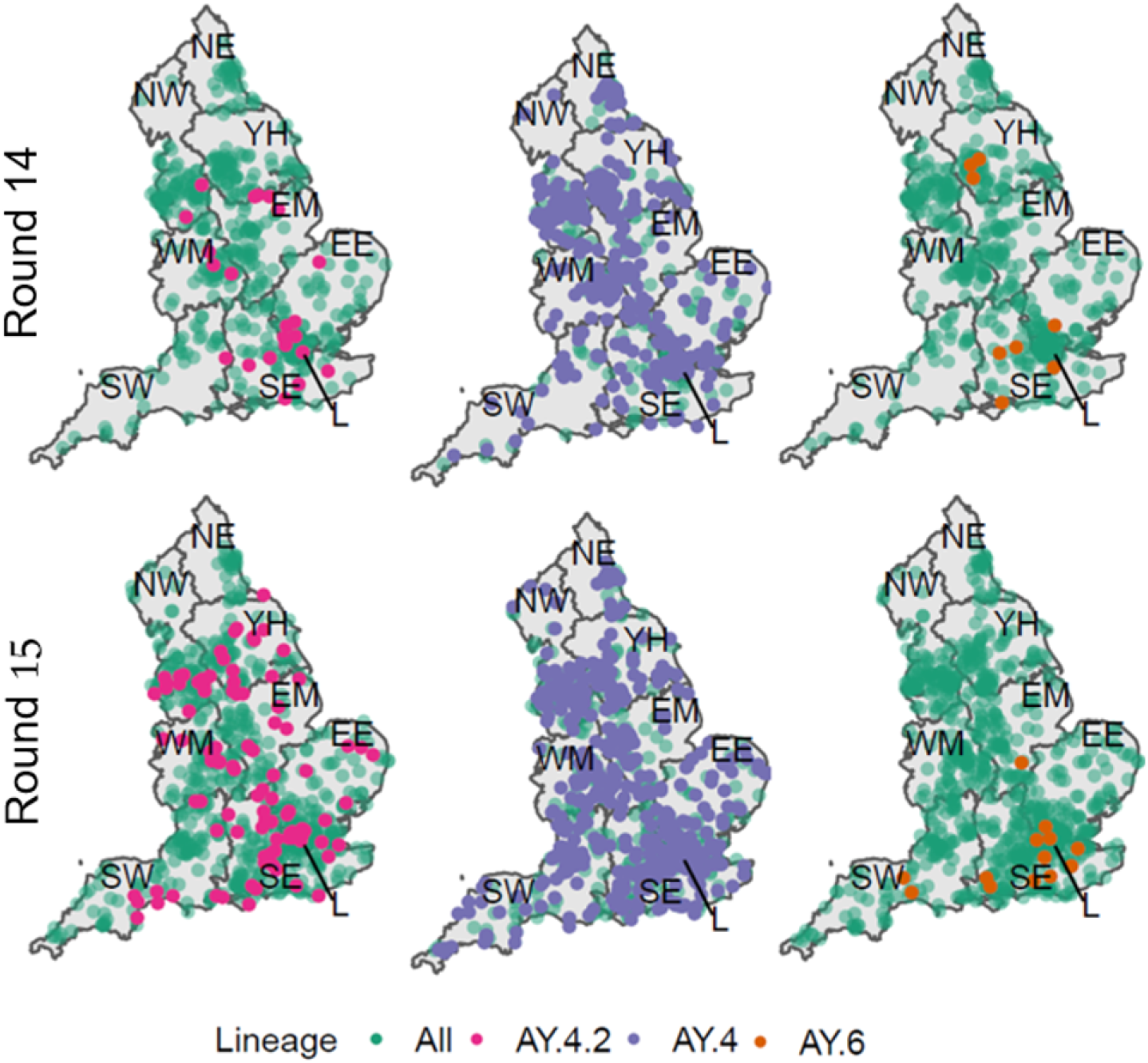
Geographic distribution of all positive samples with a lineage designation (Green) with overlaid distribution of AY.4.2 (Pink, left), AY.4 (Purple, centre) and AY.6 (Orange, right) for both round 14 (top) and round 15 (bottom). The lineages shown had either a significant level of clustering in round 14 (AY.4 and AY.4.2) or round 15 (AY.6).

During round 15 the proportion of B.1.617.2 was higher in individuals ages 25-34 years old at 24.2% (12.8%, 41.0%) relative to those aged 35-44 years old at 8.0% (4.1%, 15.0%) (p = 0.026) (Supplementary table 6). The proportion of AY.4 was found to be lower in 5-12 year olds at 52.1% (44.6%, 59.5%) relative to 35-44 year olds in which the proportion of AY.4 was 65.0% (55.3%, 73.6%) (p= 0.042) in round 15, while it was not in round 14.There were no differences between age groups in the proportion of AY.4.2 during round 15. Differences between age groups during round 14 for AY.4.2 and other lineages could not be investigated due to small sample sizes but numbers are provided in Supplementary Table 7.

### Detection of increasing sub-lineages

Logistic regression models were fitted to the proportion of each lineage detected in either round 14 or 15, allowing daily growth rates in proportion to be estimated (Figure 3, Supplementary Table 8). Of the 44 unique lineages detected, 6 were estimated to have growth rates different to zero. AY.4, AY.39, AY.98.1 and AY.111 were decreasing in proportion, whereas AY.4.2 and B.1.617.2 were increasing in proportion. The decrease in proportion of AY.4 corresponded to a daily growth rate of -0.009 (−0.015, -0.003). The increase in proportions of B.1.617.2 and AY.4.2 corresponded to growth rates of 0.029 (0.017, 0.041) and 0.028 (0.016, 0.041) respectively.

**Figure 3:**
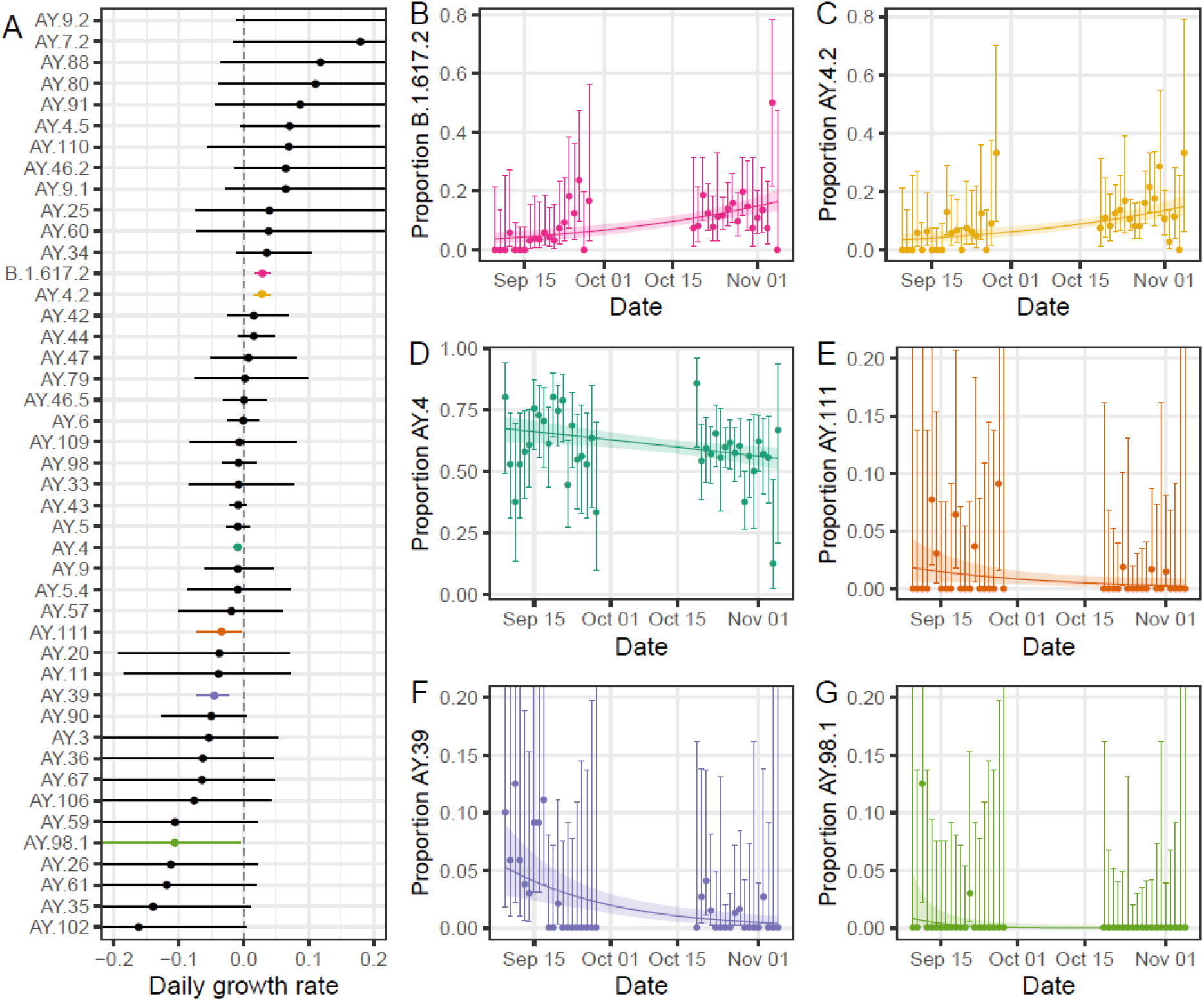
(A) Estimated daily growth rate of the log odds of each lineage detected relative to all other lineages. Shown are both lineages with a growth rate not significantly different to zero (black) and those with a growth rate significantly different to zero (coloured). (B-G) Raw estimates of the daily proportions (points) with 95% confidence intervals (error bars) for lineages with a growth rate significantly different to zero: B.1.617.2 (B, pink), AY.4.2 (C, yellow), AY.4 (D, dark green), AY.111 (E, orange), AY.39 (F, purple), AY.98.1 (G, light green). Also shown is the best-fit Bayesian logistic regression model with central estimate (solid line) and 95% credible interval (shaded region).

Comparing estimates of the reproduction number R from round 14 to round 15 for AY.4 and AY.4.2 (see Methods) we estimate a multiplicative R advantage of 1.15 (1.08, 1.23), assuming no change in the generation time distribution.

### Differences in cycle threshold values

There were quantitative differences between lineages in the N- and E-gene Ct values. The mean N- and E-gene Ct values were lowest for AY.6 though not materially lower than the values obtained for AY.4 (Figure 4, Supplementary Table 9). Mean N-gene Ct value was 22.14 (20.30, 23.99) for AY.6 compared to 23.98 (23.68, 24.28) for AY.4 (p = 0.054). Mean E-gene Ct value was 20.74 (18.90, 22.59) for AY.6 compared to 22.46 (22.16, 22.76) for AY.4 (p= 0.071). Mean N- and E-gene Ct values were found to be comparable to AY.4 for both AY.4.2 and AY.5. Relative to AY.4, mean N- and E-gene Ct values for AY.43, AY.44, AY.39 and B.1.617.2 were all higher.

**Figure 4:**
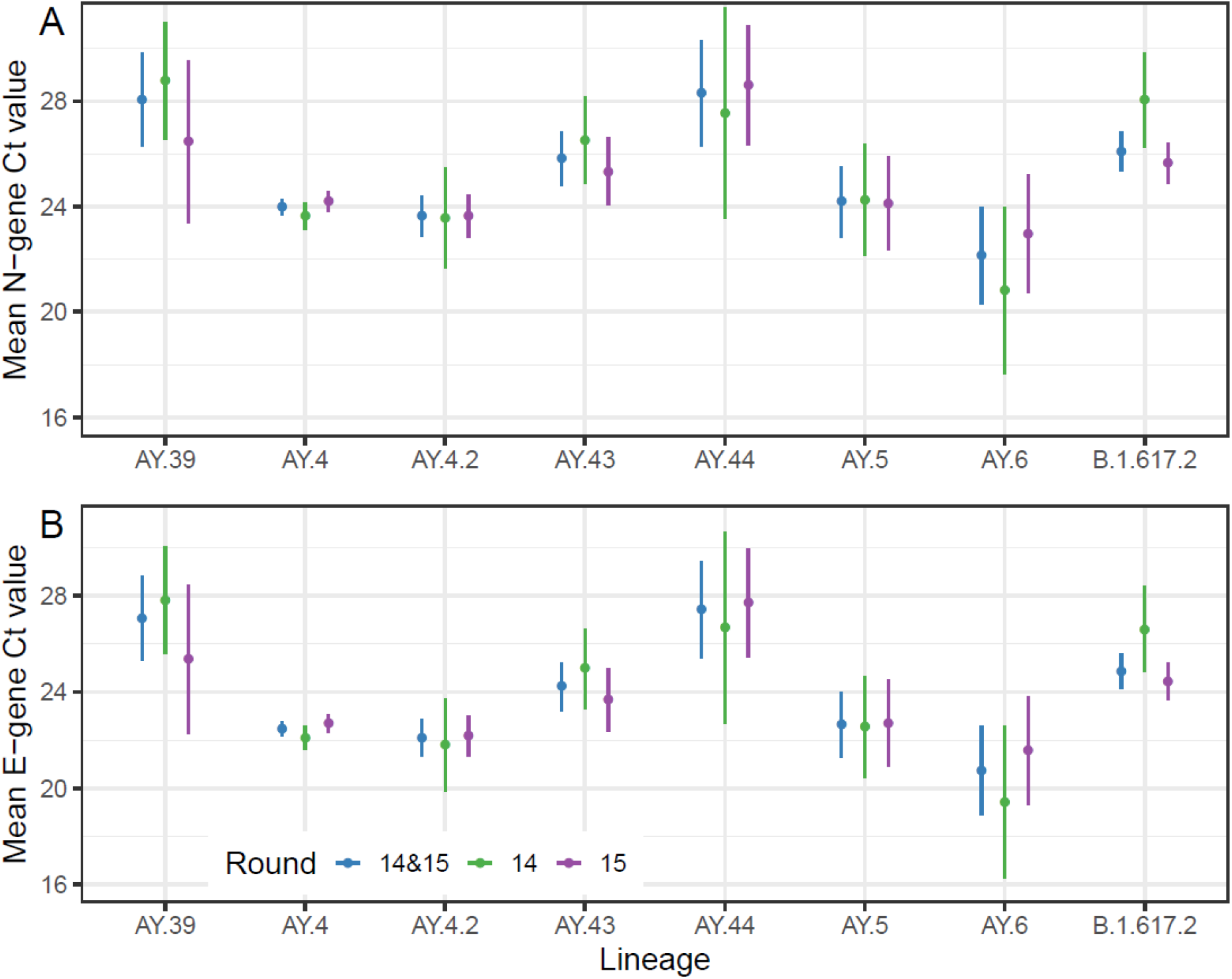
Estimated mean N-gene (A) and E-gene (B) Ct values for the 8 lineages most prevalent over rounds 14 and 15 (AY.39, AY.4, AY.4.2, AY.43, AY.44, AY.5, AY.6 and B.1.617.2) as calculated using Gaussian regression. Point estimates (points) and 95% confidence intervals (lines) are shown for estimates obtained using data from both rounds (blue), data from just round 14 (green) and data from just round 15 (purple)

### Differences in symptomatology

The proportion of individuals exhibiting the most predictive COVID-19 symptoms (loss or change of sense of taste, loss or change of sense of smell, new persistent cough, fever) in the month prior to swabbing was lower (p=0.026) in those infected with AY.4.2 at 38.1% (29.9%, 47.1%) relative to those infected with AY.4 at 49.2% (45.7%, 52.8%) (Figure 5-A, Supplementary Table 10). This difference was not explained by patterns in age or N-gene Ct value (Figure 5-B, Supplementary Table 11). In all logistic models for symptom status adjusting for round of the study, the point estimate of the effect of AY.4.2 infection was broadly similar, but, due to increased variability in the estimate, did not reach statistical significance. In particular, the odds ratio of AY.4.2-infected participants exhibiting the most predictive COVID-19 symptoms was 0.69 (0.46, 1.03) lower than for AY.4 infected participants (p= 0.069) when including round as a covariate (Model 3, Supplementary Table 11).

**Figure 5:**
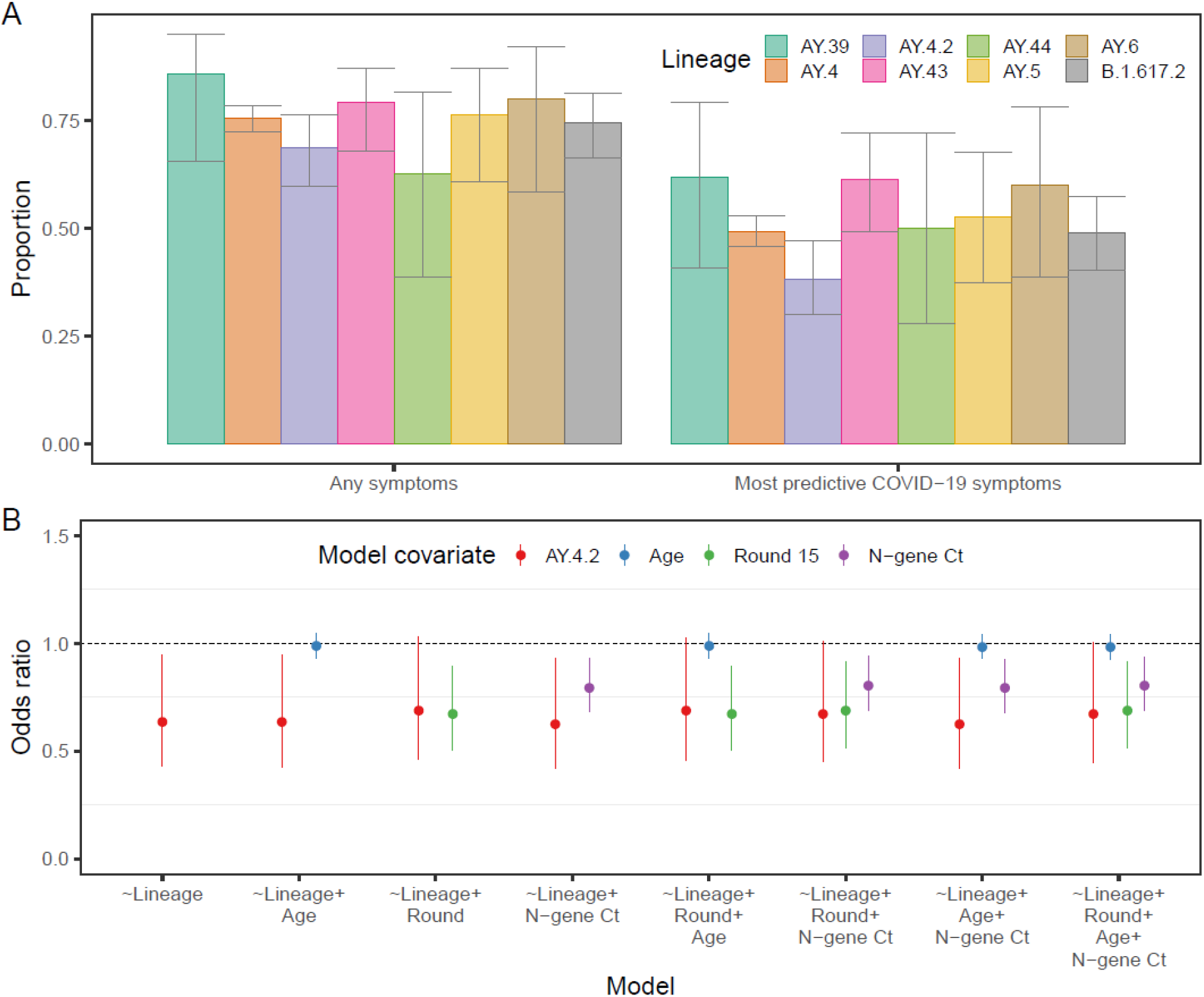
(A) Proportion of positive individuals reporting any symptoms or reporting one of the four most predictive COVID-19 symptoms (loss or change of sense of taste, loss or change of sense of smell, new persistent cough, fever) in the last month by lineage of infection, for the 8 lineages most prevalent during rounds 14 and 15 (AY.39, AY.4, AY.4.2, AY.43, AY.44, AY.5, AY.6 and B.1.617.2). (B) Odds ratios of reporting the most predictive COVID-19 symptoms in the last months for multivariable logistic regression models including lineage (AY.4.2 with reference AY.4, red), age (relative to change of 10 years in age, blue), round of study (round 15 with reference round 14, green) and N-gene Ct value (relative to change in Ct value of 5, purple).

In addition, 68.6% (59.8%, 76.3%) of those infected with AY.4.2 reported any symptoms in the month prior to swabbing compared to 75.4% (72.2%, 78.3%) for those infected with AY.4 (p= 0.119). There were no differences evident in symptom reporting between AY.4 infected individuals and the other 6 most prevalent lineages (B.1.617.2, AY.5, AY.6, AY.43, AY.44 and AY.39).

### Phylogeographic analysis

A relaxed molecular clock model was fit to the data and used to estimate a time-resolved phylogenetic tree (Figure 6). AY.4.2 was found to populate two closely related clades that emerged in June/July 2021. AY.43, AY.5 and AY.6 were also observed to have distinct clade groupings having emerged around June/July 2021 as well. The mutation rates inferred at the tree’s tips showed a large degree of variation in all of the 8 most prevalent lineages. The mean mutation rate for AY.4.2 was found to be 0.57 (<0.01, 1.10) × 10^−4^ lower than the mean mutation rate of AY.4 (p = 0.050) (Figure 6, Supplementary Table 12). The mean mutation rate inferred for samples collected in round 15 was found to be 1.00 (0.70, 1.40) × 10^−4^ lower than the mean mutation rate for samples collected in round 14 (p < 0.001).

**Figure 6:**
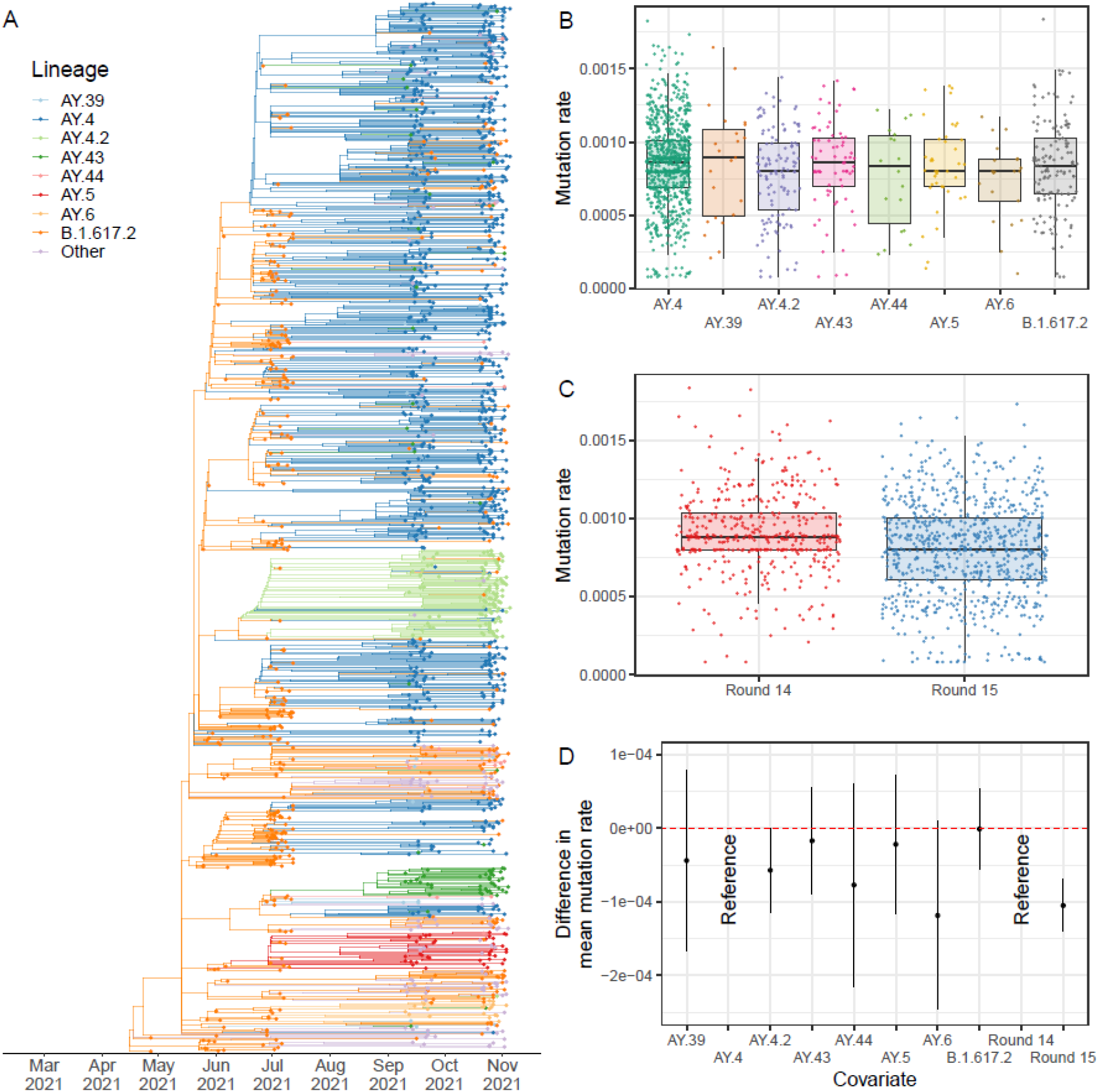
(A) Time-resolved phylogenetic tree of all positive samples obtained for which the lineage designated was Delta or a Delta sub-lineage. (B) Distribution of mutation rates inferred at each phylogenetic tree tip for the 8 lineages most prevalent in round 14 and round 15 for samples obtained in round 14 and 15. (C) Distribution of mutation rates inferred at each phylogenetic tree tip for samples collected in round 14 and round 15. (D) Difference in the mean mutation rates between lineages and rounds as inferred from a Gaussian regression model.

A mugenic model was run on the time-resolved phylogenetic tree to estimate the relative virus migration rates between regions, a measure of inter-region transmission (Supplementary Table 13). Overall levels of inter-region transmission were lowest for the North East during round 14 and 15. The highest overall level of inter-region transmission was observed for the North West during round 14 and 15, but looking at individual rounds there were higher levels for Yorkshire and The Humber in round 14 and for the South East in round 15. High rates of transmission during round 14 and 15 were found between the North West and Yorkshire and The Humber, the West Midlands and the South East, and also between the South East and London.

## Discussion

The proportion of AY.4.2 was found to be increasing between 9 September and 5 November 2021, as also reported in the routine data surveillance for England [11]. In round 15, AY.4.2 represented 11.8% of infections in line with other estimates [11]. This increase in proportion corresponded to a 15% increase in transmission advantage although this assumes the generation time distribution has remained constant; a decrease of the generation time distribution for AY.4.2 would also explain the increased growth but we are unable to test for this with prevalence data. In the past, the A222V mutation, associated with AY.4.2, increased in frequency but this was eventually deemed to be due to a founder effect and not a transmission advantage [26,27]. Given the high levels of geographic dispersion (though with some clustering) during rounds 14 and 15 it is highly unlikely that a founder effect can explain the current growth, though we can not rule out a similar effect due to higher proportions of AY.4.2 in school-aged children (prevalence increased to a greater extent in school-aged children than in adults from July to September 2021 [5,28]). However, as the proportion AY.4.2 was approximately constant by age in round 15 this growth advantage would not be detected into the future if this was the case.

Observed distributions of N- and E-gene Ct values were similar in AY.4.2 and AY.4 and so it is unlikely that the transmission advantage observed can be attributed to a higher viral load (a Ct 1 unit lower corresponds to an approximate 2-fold increase in viral load [29]). However, a reduced proportion of AY.4.2 infected individuals reporting symptoms could explain the increased transmissibility in multiple ways. Higher levels of asymptomatic infection could lead to greater levels of asymptomatic transmission. Further current testing procedures and government isolation advice in England heavily focus on the most predictive COVID-19 symptoms, which are reported less often by AY.4.2 infected individuals compared with AY.4. Thus, symptom-based policies could introduce an advantage for AY.4.2 over AY.4. Finally, the reduced level of symptom reporting could be indicative of greater levels of re-infection if AY.4.2 were more successful at evading the immune response. However, studies have found that vaccines are no less effective against AY.4.2 than other Delta sub-lineages [11] and vaccine-induced antibody neutralisation titres for AY.4.2 are similar to those for AY.4 and B.1.617.2 [30]. However, any possible evasion of the immune response caused by natural infections has yet to be investigated and the numbers reporting previous infection/vaccination is too small in this REACT-1 dataset to allow a meaningful comparison. We found a moderately reduced mutation rate of AY.4.2 relative to AY.4 which may also have introduced a fitness advantage due to a smaller number of deleterious mutations [31,32].

### Other lineages

Though we have focused on AY.4.2 we have detected a diverse set of Delta sub-lineages, with even a single detection corresponding to approximately 1000 swab-positive infections in the community at one time during the study period. The short time over which AY.4.2 went from being an undeclared lineage to a variant under investigation shows how crucial it is to have careful surveillance of all lineages irrespective of frequency. For 38 of the 44 detected lineages, it was unable to be determined whether the proportion was increasing or decreasing.

Between rounds 14 and 15 a reduction in the mean mutation rate of the virus was detected suggesting a reduction in the rate of evolution. However, despite this slowdown evolution is still occurring and we observed an increase in the proportion of B.1.617.2, an indicator that the number of undeclared B.1.617.2 sub-lineages was increasing, suggesting even further diversity of Delta sub-lineages that have yet to be given a unique lineage designation.

Further, though we capture the dynamics within England, SARS-CoV-2 is a global problem and new variants of concern can arise anywhere in the world and then spread through international travel. Higher proportions of B.1.617.2 were detected in London as well as higher levels of diversity; this likely reflects the role London continues to play in the introduction of international variants [33]. Within England, the North West region played a major role in the dissemination of the virus, having the greatest inferred rate of inter-region transmission.

Analysis of N- and E-gene Ct values found decreased levels in AY.4 and AY.4.2, which is unsurprising given both have successfully disseminated across the country, but AY.5 and AY.6 were also found to have similarly low Ct values suggesting similar viral loads; the mean N- and E-gene Ct value appeared slightly lower for AY.6 compared to AY.4. Clustering was also detected in round 15 for AY.6; careful consideration of AY.6 should be given in the future in case the current lack of growth so far reported [11] has only been due to its geographic isolation.

### Limitations

We have presented the inferred dynamics between Delta sub-lineages in England between 9 September and 5 November 2021. Our sample’s main strength over those obtained from routine surveillance is the random nature of the testing program leading to a relatively unbiased set of positive samples. However, as the sample sizes we obtain are relatively small compared with routine national surveillance our estimates have lower precision.

Lineages were only successfully determined for ∼61% of positive samples, with the ability to determine a lineage heavily influenced by a sample’s Ct value; this has potentially led to biases with lineages with lower Ct values more heavily represented in the dataset. Detecting distinct sub-lineages is a high-dimensional problem, with often many common mutations being shared between distinct lineages with only a small number of distinguishing mutations. This is exacerbated when all the lineages are highly related, as in the current nature of the pandemic in England where all samples are descendants of Delta (B.1.617.2), and can lead to incorrect designations [23]. Further, only sub-lineages that have been defined are able to be assigned to a sample. During the emergence of a new sub-lineage there is a phase of ambiguity when numbers are small and it is unclear if the mutations present warrant the declaration of a new sub-lineage. This can be seen in the detection of AY.4.2 and AY.43; both lineages had been circulating for months by October 2021 [11] but were not yet declared sub-lineages by pangoLEARN [24] in early October 2021, and so did not appear in the publicly available technical briefings [25].

## Concluding remarks

Since the beginning of the pandemic, selective pressure has led to rapid evolution in the spike protein [34] driving leaps in transmissibility [6]. However, as a greater proportion of the population acquires immunity through either infection or vaccination there will be a shift in evolutionary pressure towards immune escape. At the point of endemicity it is probable that adaptive evolution would more closely resemble the continual antigenic drift observed in influenza H3N2 [35,36]. As the evolutionary phase of SARS-CoV-2 progresses towards endemicity, continued surveillance is paramount in not only detecting increased levels of transmissibility for specific lineages, but in also better characterising the mechanism behind such changes and informing policy around testing (including case definitions).

Representative community studies such as REACT-1 can be useful in measuring the relative growth of lineages and in characterising differences in viral loads, symptomatology and geographic distribution.

## Material and methods

### Viral genome sequencing

The methods of the REACT-1 study have been described elsewhere [37]. REACT-1 is a repeat cross-sectional study whereby in each round a random subset of the English population (selected from the National Health Service general practitioners’ patient list) is invited to obtain a self-administered swab test (parent/guardian administered for 5-12 year olds). These tests are then sent to a laboratory to undergo rt-PCR testing for the presence of SARS-CoV-2. A round of the study covers a ∼2- to 3-week period and has occurred approximately monthly since May 2020 with between 100,000 and 185,000 individuals taking part in each round. Since round 8 in January 2021 all positive samples with a low enough N-gene Ct value (the threshold was 34 in rounds 14 and 15 presented here) and sufficient volume have been sent for genome sequencing. Amplification of the extracted RNA was performed using the ARTIC protocol [38] (version 4 primers), with sequence libraries prepared using CoronaHiT [39]; sequencing was performed on the Illumina NextSeq 500 platform. Raw sequences were analysed using the bioinformatic pipeline [40] and then uploaded to CLIMB [41]. Lineages were assigned using PangoLEARN [24] (database version 2021-11-04), a machine learning-based assignment algorithm, using Pango nomenclature [9]. For some sequences of low overall quality, a lineage designation was not possible and so they were not included in the analyses. Samples with less than 50% of bases covered were further excluded from the analysis.

### Phylogeographic model

For all sequences from REACT-1 rounds 11 (15 April - 3 May 2021), 12 (20 May - 7 June 2021), 13 (24 June - 12 July 2021), 14 (9 September - 27 September 2021) and 15 (19 October - 5 November 2021), in which the lineage designated was Delta or a Delta sub-lineage, a maximum likelihood phylogenetic tree was constructed using a HKY model implemented in IQ-TREE [42]. An uncorrelated relaxed clock model implemented in TreeTime [43], assuming a normal distribution of rates with mean 0.0008 substitutions per site per year and a single coalescent rate for the time scale, was then fit to the maximum likelihood phylogenetic tree producing a time-resolved phylogenetic tree. The mutation rates at the tree’s tips were extracted from the model and a Gaussian regression model was fit to the samples obtained during round 14 and 15 for the 8 most prevalent lineages (AY.39, AY.4, AY.4.2, AY.43, AY.44, AY.5, AY.6, B.617.2) including lineage and round as covariates. A mugenic model (implemented in TreeTime [43]) was run on the time-resolved phylogenetic tree, treating the region in which each sample was isolated as a discrete state. This allowed estimates of the migration rates between regions to be calculated (assumed to be symmetric).

### Statistical analyses

The 95% confidence intervals for lineage proportions were calculated using the Wilson method [44] assuming a Binomial distribution. This method is preferred when the number of positives is low but is still valid when this is not the case [45]. Higher accuracy in confidence interval estimates for when the number of positives is low was chosen so that lower bounds on case numbers for rarer lineages were as accurate as possible.

Estimates of the true number of swab-positive infections in England during round 14 and round 15 for lineages in which only one sample was detected in a round were calculated by multiplying the estimated proportion of the lineage for each round, the weighted prevalence estimated for each round [46], and the population size of England [47].

The significance of differences in proportions of particular lineages by age group and region was calculated using Fisher’s exact test with a binary outcome variable (lineage of interest or not). Differences with a p-value less than 0.05 were considered statistically significant. Analysis was only completed for a lineage in a round if there were more than 90 samples (AY.4 round 14, AY.4 round 15, AY.4.2 round 15, B.1.617.2 round 15), so that there were, on average, more than 10 samples per parameter (9 regions in England).

Shannon diversity was calculated using all data for round 14 and round 15, and for each region for round 14 and round 15 [48]. The significance of any differences in Shannon diversity between round 14 and 15 (for all data) and between regions in each round was assessed using the Hutcheson T-test [49] and its associated p-value.

The relative growth rate of a lineage compared to all other lineages was estimated using a Bayesian logistic regression model fit to the binary outcome variable (lineage of interest or not) over time. The probability that the growth rate was greater than zero was calculated from the model’s posterior. Lineages were deemed to be different to zero if the posterior probability that the growth rate was greater than zero was greater than 0.975 or less than 0.025, similar to a p-value threshold of 0.05.

The growth rates of AY.4.2 and AY.4 infected individuals were estimated by fitting an exponential model to the daily weighted prevalence using all REACT-1 data (all negatives and all AY.4/AY.4.2 associated positives) for rounds 14 and 15 assuming a Binomial likelihood. Growth rates were then converted to estimates of the reproduction number R assuming a gamma-distributed generation time with the shape parameter, n =2.29, and rate parameter, b =0.36 [50] through the equation 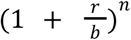 [51]. The multiplicative R advantage of AY.4.2 over AY.4 was estimated using the entire posterior distribution of *R*_*AY*.4.2_/*R*_*AY*.4_ with the median and 95% credible interval reported.

For each lineage with more than 1 sample in a round the presence of clustering was assessed. The pairwise distance matrix between all n samples that were designated to a specific lineage was calculated and from this a mean pairwise distance was calculated for the lineage. Next, 10,000 random combinations of n positive individuals (n positive individuals chosen each time without replacement), for which any lineage was determined, were selected and for each combination the distance matrix and mean distance was calculated. The proportion of the 10,000 estimated mean distances below the lineage-specific mean distance was then calculated. Clustering was deemed to be significant if this proportion was less than 0.05.

For the 8 most prevalent lineages across rounds 14 and 15 Gaussian regression was performed to estimate the mean N- and E-gene Ct values for each lineage and p-values used to assess the significance of any difference to the reference lineage (AY.4). Models were run on all data (rounds 14 and 15 combined) and then run on data from each individual round as a sensitivity analysis.

The proportion of individuals reporting any symptoms in the month prior to swabbing and any of the most predictive COVID-19 symptoms in the month prior to swabbing was calculated for the 8 most prevalent lineages across rounds 14 and 15. P-values were estimated for each lineage relative to AY.4 by performing logistic regression with the symptom status as a binary variable (any symptoms vs no symptoms, and separately most predictive COVID-19 symptoms vs none of the most predictive COVID-19 symptoms). The sensitivity of the results that AY.4.2 is less likely to exhibit the most predictive COVID-19 symptoms, relative to AY.4, was assessed by fitting further logistic regression models including age, round of study and N-gene Ct value as covariates (E-gene was also investigated but was no different to using N-gene and so this was not included).

## Data Availability

Access to REACT-1 data is restricted due to ethical and security considerations. Summary statistics and descriptive tables from the current REACT-1 study are available in the Supplementary Information. Additional summary statistics and results from the REACT-1 programme are also available at https://www.imperial.ac.uk/medicine/research-and-impact/groups/react-study/real-time-assessment-of-community-transmission-findings/ and https://github.com/mrc-ide/reactidd/tree/master/inst/extdata REACT-1 Study Materials are available for each round at https://www.imperial.ac.uk/medicine/research-and-impact/groups/react-study/react-1-study-materials/
Sequence read data are available without restriction from the European Nucleotide Archive at https://www.ebi.ac.uk/ena/browser/view/PRJEB37886, and consensus genome sequences are available from the Global initiative on sharing all influenza data at https://www.gisaid.org.

## Supplementary Tables

**Supplementary Table 1:** Lineages detected in rounds 14 and 15 of REACT-1. Supplementary Table 1 is available in this spreadsheet

**Supplementary Table 2:** Estimates of Shannon diversity for England, and by region for rounds 14 and 15 of REACT-1

Supplementary Table 2 is available in this spreadsheet

**Supplementary Table 3:** Regional distribution of AY.4 (round 14 and round 15), AY.4.2 (round 15) and B.1.617.2 (round 15).

Supplementary Table 3 is available in this spreadsheet

**Supplementary Table 4:** Raw numbers of all lineages by region for round 14 and 15 of REACT-1

Supplementary Table 4 is available in this spreadsheet

**Supplementary Table 5:** Estimated P-value for the presence of clustering for all lineages with more than a single sample in an individual round, for round 14 and 15 of REACT-1

Supplementary Table 5 is available in this spreadsheet

**Supplementary Table 6:** Distribution of AY.4 (round 14 and round 15), AY.4.2 (round 15) and B.1.617.2 (round 15) by age group.

Supplementary Table 6 is available in this spreadsheet

**Supplementary Table 7:** Raw numbers of all lineages by age group for round 14 and 15 of REACT-1

Supplementary Table 7 is available in this spreadsheet

**Supplementary Table 8:** Estimated growth rate in the log odds of every lineage detected relative to all other lineages from round 14 to 15 of REACT-1.

Supplementary Table 8 is available in this spreadsheet

**Supplementary Table 9:** Mean N- and E-gene Ct value for the eight most prevalent lineages as inferred from Gaussian regression

Supplementary Table 9 is available in this spreadsheet

**Supplementary Table 10:** Symptom status by lineage for the eight most prevalent lineages in rounds 14 and 15 of REACT-1

Supplementary Table 10 is available in this spreadsheet

**Supplementary Table 11:** Multivariable logistic regression models to determine the effect of the lineage AY.4.2 on the odds of an individual reporting any of the most predictive

COVID-19 symptoms relative to AY.4

Supplementary Table 11 is available in this spreadsheet

**Supplementary Table 12:** Multivariable gaussian regression model to determine the effect of lineage and round of the study on mean mutation rate

Supplementary Table 12 is available in this spreadsheet

**Supplementary Table 13:** Average inter-region migration rate, inferred from a mugenic model run on a time-resolved phylogenetic tree, for the periods of rounds 14 and 15, round 14 and round 15

Supplementary Table 13 is available in this spreadsheet

## Ethics

We obtained research ethics approval from the South Central-Berkshire B Research Ethics Committee (IRAS ID: 283787).

## Declaration of interests

We declare no competing interests.

## Funding

The study was funded by the Department of Health and Social Care in England. Sequencing was provided through the COVID-19 Genomics UK Consortium (COG-UK) which is supported by funding from the Medical Research Council (MRC) part of UK Research & Innovation (UKRI), the National Institute of Health Research (NIHR) [grant code: MC_PC_19027], and Genome Research Limited, operating as the Wellcome Sanger Institute.

## Acknowledgements

MC-H acknowledges support from the H2020-EXPANSE project (Horizon 2020 grant No 874627). MC-H and BB acknowledge support from Cancer Research UK, Population Research Committee Project grant ‘Mechanomics’ (grant No 22184 to MC-H). CAD acknowledges support from the MRC Centre for Global Infectious Disease Analysis and National Institute for Health Research (NIHR) Health Protection Research Unit (HPRU). GC is supported by an NIHR Professorship. HW acknowledges support from an NIHR Senior Investigator Award and the Wellcome Trust (205456/Z/16/Z). PE is Director of the Medical Research Council (MRC) Centre for Environment and Health (MR/L01341X/1, MR/S019669/1). PE acknowledges support from Health Data Research UK (HDR UK); the NIHR Imperial Biomedical Research Centre; NIHR Health Protection Research Units in Chemical and Radiation Threats and Hazards, and Environmental Exposures and Health; the British Heart Foundation Centre for Research Excellence at Imperial College London (RE/18/4/34215); and the UK Dementia Research Institute at Imperial College London (MC_PC_17114). AJP acknowledges the support of the Biotechnology and Biological Sciences Research Council (BB/R012504/1). We thank The Huo Family Foundation for their support of our work on COVID-19.

We thank key collaborators on this work – Ipsos MORI: Kelly Beaver, Sam Clemens, Gary Welch, Nicholas Gilby, Kelly Ward, Galini Pantelidou and Kevin Pickering; Institute of Global Health Innovation at Imperial College London: Gianluca Fontana, Justine Alford; School of Public Health, Imperial College London: Eric Johnson, Rob Elliott, Graham Blakoe; Quadram Institute, Norwich, UK: Alexander J. Trotter; North West London Pathology and Public Health England (now UKHSA) for help in calibration of the laboratory analyses; Patient Experience Research Centre at Imperial College London and the REACT Public Advisory Panel; NHS Digital for access to the NHS register; the Department of Health and Social Care for logistic support.

## Data availability

Access to REACT-1 data is restricted due to ethical and security considerations. Summary statistics and descriptive tables from the current REACT-1 study are available in the Supplementary Information. Additional summary statistics and results from the REACT-1 programme are also available at https://www.imperial.ac.uk/medicine/research-and-impact/groups/react-study/real-time-assessment-of-community-transmission-findings/ and https://github.com/mrc-ide/reactidd/tree/master/inst/extdata REACT-1 Study Materials are available for each round at https://www.imperial.ac.uk/medicine/research-and-impact/groups/react-study/react-1-study-materials/

Sequence read data are available without restriction from the European Nucleotide Archive at https://www.ebi.ac.uk/ena/browser/view/PRJEB37886, and consensus genome sequences are available from the Global initiative on sharing all influenza data at https://www.gisaid.org.

## Additional information

Full list of COG-UK author’s names and affiliations are available [here](https://docs.google.com/document/d/1UrDM9fLyZ8aGoN0M6hcd9863Ox6_IZbB/edit)

## Notes

### Competing Interest Statement

The authors have declared no competing interest.

## References

1. Lineage B.1.617.2 Pangolin report. [cited 26 Nov 2021]. Available: https://cov-lineages.org/global_report_B.1.617.2.html

2. World Health Organisation. COVID-19 Weekly Epidemiological Update - Edition 66. 2021. [cited 23 Nov 2021]. Available: https://www.who.int/publications/m/item/weekly-epidemiological-update-on-covid-19---16-november-2021

3. Bernal JL, Andrews N, Gower C, Gallagher E, Simmons R, Thelwall S, et al. Effectiveness of COVID-19 vaccines against the B.1.617.2 variant. medRxiv. 2021; 2021.05.22.21257658.

4. PHE Genomics Cell, PHE Outbreak Surveillance Team, PHE Epidemiology Cell, PHE Contact Tracing Data Team, PHE Health, Protection Data Science Team, PHE Joint Modelling Team, NHS Test and Trace Joint Biosecurity Centre, Public Health Scotland and EAVE group, Contributions from the Variant Technical Group Members. SARS-CoV-2 variants of concern and variants under investigation in England - Technical briefing 15, 11 June 2021. Available: https://assets.publishing.service.gov.uk/government/uploads/system/uploads/attachment_data/file/993879/Variants_of_Concern_VOC_Technical_Briefing_15.pdf

5. Elliott P, Haw D, Wang H, Eales O, Walters CE, Ainslie KEC, et al. Exponential growth, high prevalence of SARS-CoV-2, and vaccine effectiveness associated with the Delta variant. Science. 2021; eabl9551.

6. Obermeyer F, Schaffner SF, Jankowiak M, Barkas N, Pyle JD, Park DJ, et al. Analysis of 2.1 million SARS-CoV-2 genomes identifies mutations associated with transmissibility. doi:10.1101/2021.09.07.21263228

7. Baj A, Novazzi F, Drago Ferrante F, Genoni A, Tettamanzi E, Catanoso G, et al. Spike protein evolution in the SARS-CoV-2 Delta variant of concern: a case series from Northern Lombardy. Emerg Microbes Infect. 2021;10: 2010–2015.

8. Kistler KE, Huddleston J, Bedford T. Rapid and parallel adaptive mutations in spike S1 drive clade success in SARS-CoV-2. bioRxiv. 2021. doi:10.1101/2021.09.11.459844

9. Rambaut A, Holmes EC, O’Toole Á, Hill V, McCrone JT, Ruis C, et al. A dynamic nomenclature proposal for SARS-CoV-2 lineages to assist genomic epidemiology. Nat Microbiol. 2020;5: 1403–1407.

10. UKHSA Genomics Cell UKHSA Outbreak Surveillance Team UKHSA Epidemiology Cell UKHSA Contact Tracing Data Team UKHSA International Cell UKHSA Environmental Monitoring for Health Protection Team. SARS-CoV-2 variants of concern and variants under investigation in England - Technical briefing 27, 29 October 2021. Available: https://assets.publishing.service.gov.uk/government/uploads/system/uploads/attachment_data/file/1029715/technical-briefing-27.pdf

11. UKHSA Genomics Cell UKHSA Outbreak Surveillance Team UKHSA Epidemiology Cell UKHSA Contact Tracing Data Team UKHSA International Cell UKHSA Environmental Monitoring for Health Protection Team. SARS-CoV-2 variants of concern and variants under investigation in England - Technical briefing 28, 12 November 2021. Available: https://assets.publishing.service.gov.uk/government/uploads/system/uploads/attachment_data/file/1033101/Technical_Briefing_28_12_Nov_2021.pdf

12. UKHSA Genomics Cell UKHSA Outbreak Surveillance Team UKHSA Epidemiology Cell UKHSA Contact Tracing Data Team UKHSA International Cell UKHSA Environmental Monitoring for Health Protection Team. SARS-CoV-2 variants of concern and variants under investigation in England - Technical briefing 26, 22 October 2021. Available: https://assets.publishing.service.gov.uk/government/uploads/system/uploads/attachment_data/file/1028113/Technical_Briefing_26.pdf

13. Lineage AY.4.2 Pangolin report. [cited 23 Nov 2021]. Available: https://cov-lineages.org/lineage.html?lineage=AY.4.2

14. Alaa Abdel Latif, Julia L. Mullen, Manar Alkuzweny, Ginger Tsueng, Marco Cano, Emily Haag, Jerry Zhou, Mark Zeller, Emory Hufbauer, Nate Matteson, Chunlei Wu, Kristian G. Andersen, Andrew I. Su, Karthik Gangavarapu, Laura D. Hughes, and the Center for Viral Systems Biology. AY.4 lineage report, outbreak.info. [cited 23 Nov 2021]. Available: https://outbreak.info/situation-reports?pango=AY.4.2

15. Lineage AY.4 Pangolin report. [cited 23 Nov 2021]. Available: https://cov-lineages.org/lineage.html?lineage=AY.4

16. Umair M, Ikram A, Rehman Z, Haider A, Badar N, Ammar M, et al. Genomic diversity of SARS-CoV-2 in Pakistan during fourth wave of pandemic. bioRxiv. 2021. doi:10.1101/2021.09.30.21264343

17. Danish Covid-19 Genome Consortium. Genomic overview of SARS-CoV-2 in Denmark, 19 November 2021. [cited 23 Nov 2021]. Available: https://www.covid19genomics.dk/statistics

18. Official UK Coronavirus Dashboard. [cited 17 May 2021]. Available: https://coronavirus.data.gov.uk/

19. Latest insights team. Coronavirus (COVID-19) latest insights - Office for National Statistics. Office for National Statistics; 23 Nov 2021 [cited 25 Nov 2021]. Available: https://www.ons.gov.uk/peoplepopulationandcommunity/healthandsocialcare/conditionsanddiseases/articles/coronaviruscovid19latestinsights/antibodies

20. Riley S, Ainslie KEC, Eales O, Walters CE, Wang H, Atchison C, et al. Resurgence of SARS-CoV-2: detection by community viral surveillance. Science. 2021. doi:10.1126/science.abf0874

21. Ricoca Peixoto V, Nunes C, Abrantes A. Epidemic Surveillance of Covid-19: Considering Uncertainty and Under-Ascertainment. Portuguese Journal of Public Health. 2020;38: 23–29.

22. Feng S, Phillips DJ, White T, Sayal H, Aley PK, Bibi S, et al. Correlates of protection against symptomatic and asymptomatic SARS-CoV-2 infection. Nat Med. 2021;27:2032–2040.

23. O’Toole Á, Scher E, Underwood A, Jackson B, Hill V, McCrone JT, et al. Assignment of epidemiological lineages in an emerging pandemic using the pangolin tool. Virus Evol. 2021;7:veab064.

24. Phylogenetic Assignment of Named Global Outbreak LINeages (PANGOLIN). Github; Available: https://github.com/cov-lineages/pangolin

25. UKHSA Genomics Cell UKHSA Outbreak Surveillance Team UKHSA Epidemiology Cell UKHSA Contact Tracing Data Team UKHSA International Cell UKHSA Environmental Monitoring for Health Protection Team. SARS-CoV-2 variants of concern and variants under investigation in England - Technical briefing 25, 1 October 2021. Available: https://assets.publishing.service.gov.uk/government/uploads/system/uploads/attachment_data/file/1025827/Technical_Briefing_25.pdf

26. Díez-Fuertes F, Iglesias-Caballero M, García-Pérez J, Monzón S, Jiménez P, Varona S, et al. A Founder Effect Led Early SARS-CoV-2 Transmission in Spain. J Virol. 2021;95. doi:10.1128/JVI.01583-20

27. Hodcroft EB, Zuber M, Nadeau S, Vaughan TG, Crawford KHD, Althaus CL, et al. Spread of a SARS-CoV-2 variant through Europe in the summer of 2020. Nature. 2021;595: 707–712.

28. Chadeau-Hyam M, Wang H, Eales O, Haw D, Bodinier B, Whitaker M, et al. REACT-1 study round 14: High and increasing prevalence of SARS-CoV-2 infection among school-aged children during September 2021 and vaccine effectiveness against infection in England. medRxiv. 2021; 2021.10.14.21264965.

29. Yelin I, Aharony N, Tamar ES, Argoetti A, Messer E, Berenbaum D, et al. Evaluation of COVID-19 RT-qPCR Test in Multi sample Pools. Clin Infect Dis. 2020;71: 2073–2078.

30. Lassaunière R, Polacek C, Fonager J, Bennedbæk M, Boding L, Rasmussen M, et al. Neutralisation of the SARS-CoV-2 Delta sub-lineage AY.4.2 and B.1.617.2 + E484K by BNT162b2 mRNA vaccine-elicited sera. bioRxiv. 2021. doi:10.1101/2021.11.08.21266075

31. Peck KM, Lauring AS. Complexities of Viral Mutation Rates. J Virol. 2018;92. doi:10.1128/JVI.01031-17

32. Koelle K, Rasmussen DA. The effects of a deleterious mutation load on patterns of influenza A/H3N2’s antigenic evolution in humans. eLife. 2015;4: e07361.

33. Eales O, Page AJ, Tang SN, Walters CE, Wang H, Haw D, et al. SARS-CoV-2 lineage dynamics in England from January to March 2021 inferred from representative community samples. medRxiv; 2021. doi:10.1101/2021.05.08.21256867

34. Saputri DS, Li S, van Eerden FJ, Rozewicki J, Xu Z, Ismanto HS, et al. Flexible, Functional, and Familiar: Characteristics of SARS-CoV-2 Spike Protein Evolution. Front Microbiol. 2020;11: 2112.

35. Bedford T, Riley S, Barr IG, Broor S, Chadha M, Cox NJ, et al. Global circulation patterns of seasonal influenza viruses vary with antigenic drift. Nature. 2015;523: 217–220.

36. Andrew Rambaut, Oliver G. Pybus, Martha I. Nelson, Cecile Viboud, Jeffery K. Taubenberger, Edward C. Holmes. The genomic and epidemiological dynamics of human influenza A virus. Nature. 2008. Available: https://www.nature.com/articles/nature06945

37. Riley S, Atchison C, Ashby D, Donnelly CA, Barclay W, Cooke G, et al. REal-time Assessment of Community Transmission (REACT) of SARS-CoV-2 virus: Study protocol. Wellcome Open Research. 2020;5: 200.

38. Quick J. nCoV-2019 sequencing protocol v3 (LoCost). 2020 [cited 4 May 2021]. Available: https://www.protocols.io/view/ncov-2019-sequencing-protocol-v3-locost-bh42j8ye

39. Baker DJ, Aydin A, Le-Viet T, Kay GL, Rudder S, de Oliveira Martins L, et al. CoronaHiT: high-throughput sequencing of SARS-CoV-2 genomes. Genome Med. 2021;13: 21.

40. A Nextflow pipeline for running the ARTIC network’s field bioinformatics tools. Github; Available: https://github.com/connor-lab/ncov2019-artic-nf

41. Connor TR, Loman NJ, Thompson S, Smith A, Southgate J, Poplawski R, et al. CLIMB (the Cloud Infrastructure for Microbial Bioinformatics): an online resource for the medical microbiology community. Microb Genom. 2016;2: e000086.

42. Nguyen L-T, Schmidt HA, von Haeseler A, Minh BQ. IQ-TREE: a fast and effective stochastic algorithm for estimating maximum-likelihood phylogenies. Mol Biol Evol. 2015;32: 268–274.

43. Sagulenko P, Puller V, Neher RA. TreeTime: Maximum-likelihood phylodynamic analysis. Virus Evol. 2018;4: vex042.

44. Wilson EB. Probable Inference, the Law of Succession, and Statistical Inference. J Am Stat Assoc. 1927;22: 209–212.

45. Brown LD, Tony Cai T, DasGupta A. Interval Estimation for a Binomial Proportion. SSO Schweiz Monatsschr Zahnheilkd. 2001;16: 101–133.

46. Chadeau-Hyam M, Eales O, Bodinier B, Wang H, Haw D, Whitaker M, et al. REACT-1 round 15 final report: Increased breakthrough SARS-CoV-2 infections among adults who had received two doses of vaccine, but booster doses and first doses in children are providing important protection. 2021.[cited 26 Nov 2021]. Available: http://spiral.imperial.ac.uk/handle/10044/1/92501

47. Park N. Population estimates for the UK, England and Wales, Scotland and Northern Ireland - Office for National Statistics. Office for National Statistics; 24 Jun 2021 [cited 11 Aug 2021]. Available: https://www.ons.gov.uk/peoplepopulationandcommunity/populationandmigration/populationestimates/bulletins/annualmidyearpopulationestimates/mid2020

48. Spellerberg IF, Fedor PJ. A tribute to Claude Shannon (1916-2001) and a plea for more rigorous use of species richness, species diversity and the “Shannon-Wiener” Index. Glob Ecol Biogeogr. 2003;12: 177–179.

49. Hutcheson K. A test for comparing diversities based on the Shannon formula. J Theor Biol. 1970;29: 151–154.

50. Bi Q, Wu Y, Mei S, Ye C, Zou X, Zhang Z, et al. Epidemiology and Transmission of COVID-19 in Shenzhen China: Analysis of 391 cases and 1,286 of their close contacts. MedRxiv. 2020. Available: https://www.medrxiv.org/content/medrxiv/early/2020/03/19/2020.03.03.20028423.full.pdf

51. Wallinga J, Lipsitch M. How generation intervals shape the relationship between growth rates and reproductive numbers. Proc Biol Sci. 2007;274: 599–604.

